# Correlation of National and Healthcare Workers COVID-19 Infection Data; Implications for Large-scale Viral Testing Programs

**DOI:** 10.1101/2020.08.21.20179283

**Authors:** Dan Wu, Pól Mac Aonghusa, Donal F. O’Shea

**Author notes:** **Funding:** The authors received no specific funding for this work.

## Abstract

Time analysis of the course of an infectious disease epidemic is a critical way to understand the dynamics of pathogen transmission and the effect of population scale interventions. Computational methods have been applied to the progression of the COVID-19 outbreak in five different countries (Ireland, Germany, UK, South Korea and Iceland) using their reported daily infection data. A Gaussian convolution smoothing function constructed a continuous epidemic line profile that was segmented into longitudinal time series of mathematically fitted individual logistic curves. The time series of fitted curves allowed comparison of disease progression with differences in decreasing daily infection numbers following the epidemic peak being of specific interest. A positive relationship between rate of declining infections and countries with comprehensive COVID-19 testing regimes existed. In contrast, extended epidemic timeframes were recorded for those least prepared for large scale testing and contact tracing. As many countries continue to struggle to implement population wide testing it is prudent to explore additional measures that could be employed. Comparative analysis of healthcare worker (HCW) infection data from Ireland shows it closely related to that of the entire population with respect to trends of daily infection numbers and growth rates over a 57-day period. With 31.6% of all test-confirmed infections in healthcare workers (all employees of healthcare facilities), they represent a concentrated 3% subset of the national population which if exhaustively tested (regardless of symptom status) could provide valuable information on disease progression in the entire population (or set). Mathematically, national population and HCWs can be viewed as a set and subset with significant influences on each other, with solidarity between both an essential ingredient for ending this crisis.

## Introduction

At time of writing, the coronavirus disease 2019 (COVID-19) pandemic has passed its first peak in Europe and been suppressed in several Asian and Australasian countries, but is growing in the Americas.[1] The outbreak commenced in Wuhan, China in late 2019, with the first confirmed European cases in January associated with travel between the regions.[2,3,4] With infection numbers rising and an absence of either vaccine or proven therapeutic treatments, a series of non-pharmaceutical interventions were adopted to limit the spread of the disease.

The WHO universally advised countries to “test, test, test” with follow up contact tracing and isolation of infected individuals to contain the pandemic.[5] While this guidance is unquestionably correct, the success of its implementation has been highly variable. In parallel with this strategy, many nations required their citizens to remain in their place of residence to stunt the exponential epidemic growth phase, colloquially known as flattening the curve. This collective hibernation of national populations is a unique event in modern times, being adopted with differing levels of rigor. For example in China, infected individuals were centrally isolated in dedicated quarantine centres, Fangcang shelter hospitals, whereas self-isolation was in place of residence in European countries.[6] These drastic interventions have had a significant positive influence on reducing the rate and extent of disease progression. The opportunity now exists to examine and cross compare their outcomes such that the insights gained may assist in the advent of a second pandemic wave.[7]

In this paper, an in-depth comparative approach to “looking under the epidemic wave” is presented, using daily confirmed COVID-19 infection data from five countries - Ireland, Germany, UK, South Korea and Iceland.[8] Specifically, for Ireland, correlations between daily COVID-19 infections in the entire population and those in health care workers (HCWs) were investigated as HCW data was nationally collected from the start of the outbreak. Ireland identified its first confirmed infection on the 29^th^ of February, with schools and universities closed on March 12^th^ and a national lockdown imposed on March 27^th^. The lockdown closed most businesses and leisure amenities, restricted contact to individuals within a residence and prohibited all non-essential travel. These measures remained in place until May 18^th^ at which stage a phased relaxing of lockdown measures commenced.[9] Data analysis from the epidemic outset through to the easing of lockdown for both cohorts identified potentially useful correlations, suggesting that a HCW focused COVID-19 testing program could effectively inform of changes in national disease status.

## Methods and Data

### Data Sources

Daily confirmed COVID-19 infection data for Germany, UK, South Korea and Iceland were taken from the open COVID-19 Data Repository provided by the Center for Systems Science and Engineering (CSSE) at Johns Hopkins University.[10] The confirmed COVID-19 national and HCW infection data for Ireland was accessed from “Ireland’s COVID-19 Data Hub” published by the Government of Ireland.[11]

### Software and data processing

In brief, daily confirmed infected case data was processed in three stages. First, a moving average was calculated over a five-day window, allowing smaller windows at endpoints so that the total number of data points is preserved. Next, a Gaussian convolution filter was applied to the data for smoothing and line fitting to produce what we term the epidemic profile line (Figure 1). Finally, a series of logistic curves were fitted to the epidemic profile line making it possible to divide the epidemic timeline into growth, peak and decline stages. Logistic functions occur regularly in the study of epidemics and fitting infection data to series of logistic functions has been utilized previously.[8]

Daily confirmed COVID-19 infections from 1^st^ March to 18^th^ May 2020 for Ireland, Germany, UK, Iceland and Jan 26^th^ to April 16^th^ for South Korea were accessed using Python scripts to download and perform pre-processing stages. Five-day rolling arithmetic average of daily-confirmed infection cases were calculated during the initial pre-processing step.

### Software

PeakFit software v4.12 (Systat Software Inc) was used for generating convolved epidemic profile lines and fitting of logistic curves from averaged daily confirmed COVID-19 infection data for Ireland, Germany, UK, Iceland and South Korea (other commercial and freely available software would also be suitable). Using the autofit peaks I function, the smooth processing to produce the epidemic profile line was completed by using convolve method with 0.5% smoothing level to create a continuous smooth line from the five day rolling average data (Figure 1). Logistic curve fitting was performed to decompose epidemic profile line data into separate logistic functions. The form of logistic function used during curve-fitting is summarized in equation 1, showing the three parameters (a_0,_ a_1,_ a_2_) used. The full width at half maxima were equal for all fitted curves in any specific plot. A threshold limit of 8% of the maximum of the epidemic profile line data was taken as a lower-bound cutoff for the amplitude of each component logistic function. This value was chosen empirically to allow fitting of small peak values while providing a reasonable cutoff to prevent over-fitting with many small amplitude logistic functions. Optimum fit was achieved by repeating the software curve fitting three times for each dataset, optimizing r^2^ for goodness of fit (Figure 2). A comprehensive discussion of the processing techniques used here, including Gaussian Fourier deconvolution and curve fitting is available.[12]

**Figure 1.**
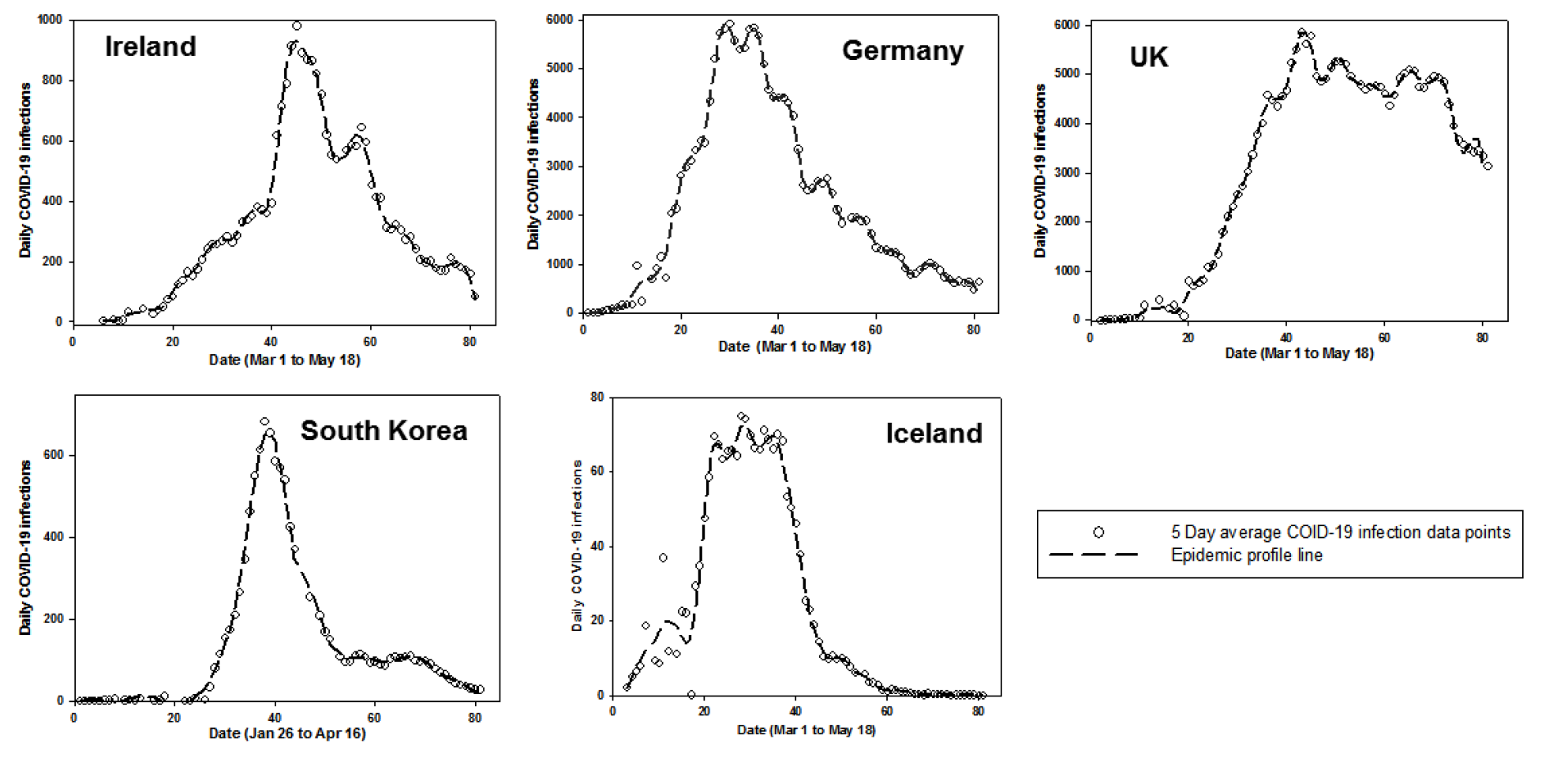
Epidemic profile lines for Ireland, Germany, UK, South Korea and Iceland. Plots of five-day averaged confirmed COVID-19 infections as a best-fit smooth epidemic profile line.

**Figure 2.**
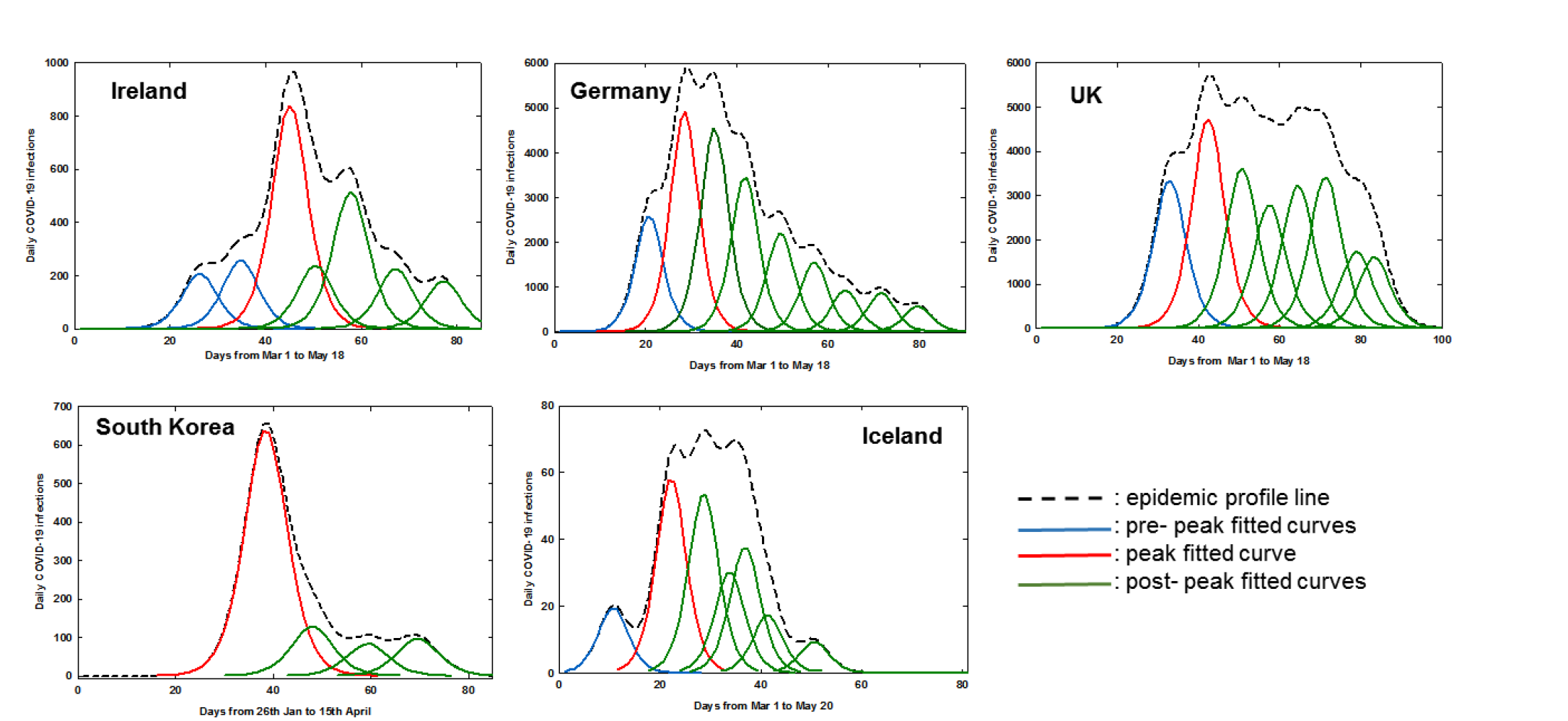
Fitted logistic curves of daily COVID-19 infections for Ireland, Germany, UK, South Korea and Iceland.

The number of days before maximum daily infections was calculated from once 0.5% of the total number of a countries COVID-19 infections was exceeded (as at this point it would be expected that community transmission was occurring) to the maxima of the fitted logistic curve of highest amplitude (shown in red in Figure 2).

**Equation 1**. Logistic function used for curve-fitting.

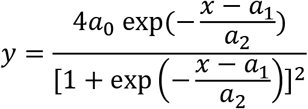

a_0_ = amplitude; a_1_ = centre value; a_2_ = width (>0)

Using the Ireland’s COVID-19 Data Hub dataset, daily percentage increase or decrease in growth rates were calculated using five day rolling averaged data using the formula (D_x_ − D_x_−1/D_x_−1)100 where Dx is the number for a specific day and Dx-1 is the number for the preceding day (Figure 7).[11] Normalization data (as shown in Figure 6) were generated by dividing each data point by the maximum valued data point to create a new data set ranged from 0 to 1. Spearman and Pearson correlation coefficients were calculated using standard methods.

## Results

From the outset, the primary focus of nations has been to limit the scale of the epidemic peak. Arguably, equally challenging is managing the descent from daily infection peak to near zero cases as quickly as possible while maintaining the balance between reopening society and not causing a surge in new infections. A method that mathematically segments the daily data over an 80-day period has been employed to illustrate how this has varied in different countries. Five Global North countries (Ireland, Germany, UK, South Korea and Iceland) of varying population and epidemic scales were selected for data cross comparison. Four of the countries chosen (Ireland, Germany, UK, and South Korea) had an accumulative number of confirmed cases in excess of 25,000 and each had, for a time, daily infection numbers between 650 and 6,300. In addition, one country with a smaller absolute numbers of infections and cumulative cases (due to smaller population), Iceland, was included to test how this mode of analysis performed for such datasets.[13]

This was achieved by first applying a smoothing convolution algorithm to the five-day rolling average infection data for each country to give a continuous epidemic profile line (Figure 1).

Next, the area under each epidemic profile line was fitted with sequential curves generated using logistic amplitude function to achieve the best coefficient of determination, r² (equation 1). Values of r^2^ between 0.976 and 0.994 were achieved for all countries (Table 1). The number of software-generated logistic curves differed for each nation’s data set and defined different phases of the epidemic progression for each country into pre-peak (blue), peak (red) and post-peak (green) segments (Figure 2). The date of fitted curve maxima were used to compare timeframes for the different national epidemics (Table 1). It is noteworthy that the lead up to the maximum number of daily cases was more gradual in Ireland and the UK relative to Germany and Iceland, with South Korea being the fastest (Figure 2, blue curves). This analysis measured Ireland and the UK at 30 and 26 days respectively, with Germany and Iceland at 18 days and South Korea at only 12 days (Table 1). Rationales for these differences could be geographical with South Korea being closest to the disease source in China and an effective TTQ implementation allowing suppression of their outbreak as it emerged. South Korea has been widely acknowledged as having a successful testing and tracing program, with valuable experienced gained from the 2003 SARS outbreak.[14] The more prolonged timeframes to reach maximum daily cases in Ireland, Germany and the UK may be associated with the disease being in wider community circulation than detected at the time of lockdown, with intra-domicile spreading of infection as quarantine took place in the home. If infected individuals existed in pre or asymptomatic stages before lockdown, once lockdown occurred they would be confined within their domiciles, taking up to weeks before newly infected individuals would show symptoms.[15]

**Table 1.** Comparison of fitted logistic curve data for different countries.

| Country | Date of >0.5% accumulative cases<sup>a</sup> | r<sup>2</sup> | Curves pre-highest amplitude | Date of highest amplitude<sup>b</sup> | Days to highest amplitude<sup>c</sup> | Curves post highest amplitude | Days from highest amplitude to 50% and (90%) reduction<sup>d</sup> |
| --- | --- | --- | --- | --- | --- | --- | --- |
| Ireland | Mar 15 | 0.988 | 2 | Apr 14 | 30 | 4 | 16 (35) |
| Germany | Mar 10 | 0.992 | 1 | Mar 28 | 18 | 7 | 21 (52) |
| UK | Mar 16 | 0.982 | 1 | Apr 11 | 26 | 6 | nr (nr)<sup>e</sup> |
| South Korea | Feb 21 | 0.994 | 0 | Mar 4 | 12 | 3 | 8 (36) |
| Iceland | Mar 4 | 0.976 | 1 | Mar 22 | 18 | 5 | 21 (34) |

<sup>a</sup> Calculated based on total number of COVID-19 confirmed cases up to May 18. <sup>b</sup> Taken as maximum of highest amplitude peak. <sup>c</sup> Calculated from date of 0.5% accumulative cases to date of highest amplitude. <sup>d</sup> Calculated based on time from peak of highest amplitude curve to 50% and 90% reduction. <sup>e</sup> Not reached (nr) by May 18. Population: Ireland 4.9 M; Germany 83.7 M; UK 67.8 M; South Korea 51.2 M; Iceland 0.36 M.

While many model systems consider the growth phase up to the epidemic maximum, analysis of the post-peak declining phase may be just as informative especially as it is closely linked with the lessening of societal restrictions. This analysis showed clear differences between the five countries studied, such as the number of post-peak logistic curves (green) and time taken to reach 50 and 90% reduction points. In an idealized scenario, once the maximum daily infections has been reached (red logistic curve) the decline in the number of new infections would follow this logistic curve to zero infections (Figure 2). Examination of the fitted green curves and key statistics shows that Ireland and Germany had similar profiles, with South Korea and Iceland being better and the UK having the poorest. In detail, the number of days from the highest amplitude curve to a 50% decrease in daily infections was 8 for South Korea whereas Ireland took 16 days, Germany and Iceland 21 and the UK > 40 days. The challenge of totally suppressing infections is revealed by the fact that from this point it took a further 19, 31, 28 or 13 days to reach a 90% reduction from maximum daily infections for Ireland, Germany, South Korea and Iceland respectively (Table 1). The analysis shows that South Korea was best able to suppress community propagation of disease and this correlates with their proficient TTQ program. In contrast, the UK experienced setbacks to their TTQ programs mid-epidemic and endured the most prolonged outbreak.[16] The quality of the logistic curve fitting for each datasets is shown utilizing 95% confidence intervals in the supporting Figures S1-S5.

Extending this mode of analysis from static datasets (as above) to dynamic datasets is also possible allowing the progress of the epidemic to be tracked in real-time. The range in which a future individual observation or next daily data point will fall can be defined using 95% predictive intervals. As each new daily data point is added, it will either coincide with the 95% predictive intervals (based on previous data points) or not. At any time point, the time leading logistic curve, if followed, maps the fastest theoretical pathway to zero infections or the end of the epidemic. Breaking of the prediction interval boundaries reveals when new points are outside the predictive intervals, causing an adjustment of curve fittings to accommodate the new data. If infection numbers do not progress to zero then an additional logistic curve would be needed to match the actual data points, indicating that epidemic is ongoing.

An example is shown below focusing on data for Ireland from April 18^th^ to 23^rd^ (Figure 3). Starting with data from Mar 1^st^ to April 17^th^ new daily data points were sequentially added, with the epidemic profile line and logistic curves re-calculated each time. On April 18^th^ the most time forward leading curve (cyan colour) had two data points (indicated by arrows) beyond the maximum of the leading logistic curve, indicating that a downward progression of infection numbers had begun. Following addition of the next data point, the leading curve remained within the 95% predictive intervals. However, on addition of the subsequent daily data point (panel C) the predictive intervals widen, indicating that the data fit is worsening. Following the addition of the daily data point for April 21^st^ (panel D) the prediction intervals break, indicating the new data does not follow the pathway projected by the leading logistic curve. It is noteworthy that each of these five daily data points were sequentially lower than the preceding one, so while a downward trajectory was occurring, the prediction intervals alerted that an underlying deviation was in progress that is not immediately obvious from the data points themselves. Following the addition of data for the next two days, a new leading logistic fitted curve (cyan coloured) is calculated and the prediction intervals come back into range. The predictive cycle can now repeat with this curve and once again this pathway is not followed by the emerging data and a similar breaking of prediction intervals occurs on April 25^th^ - 27^th^ (SI Movie 1). The entire epidemic time sequence for Ireland has been compiled into a graphical movie in which the continuum of change from evolution in the early stage of the outbreak through to containment in the later stages can be visually observed (SI Movie 1).

**Figure 3.**
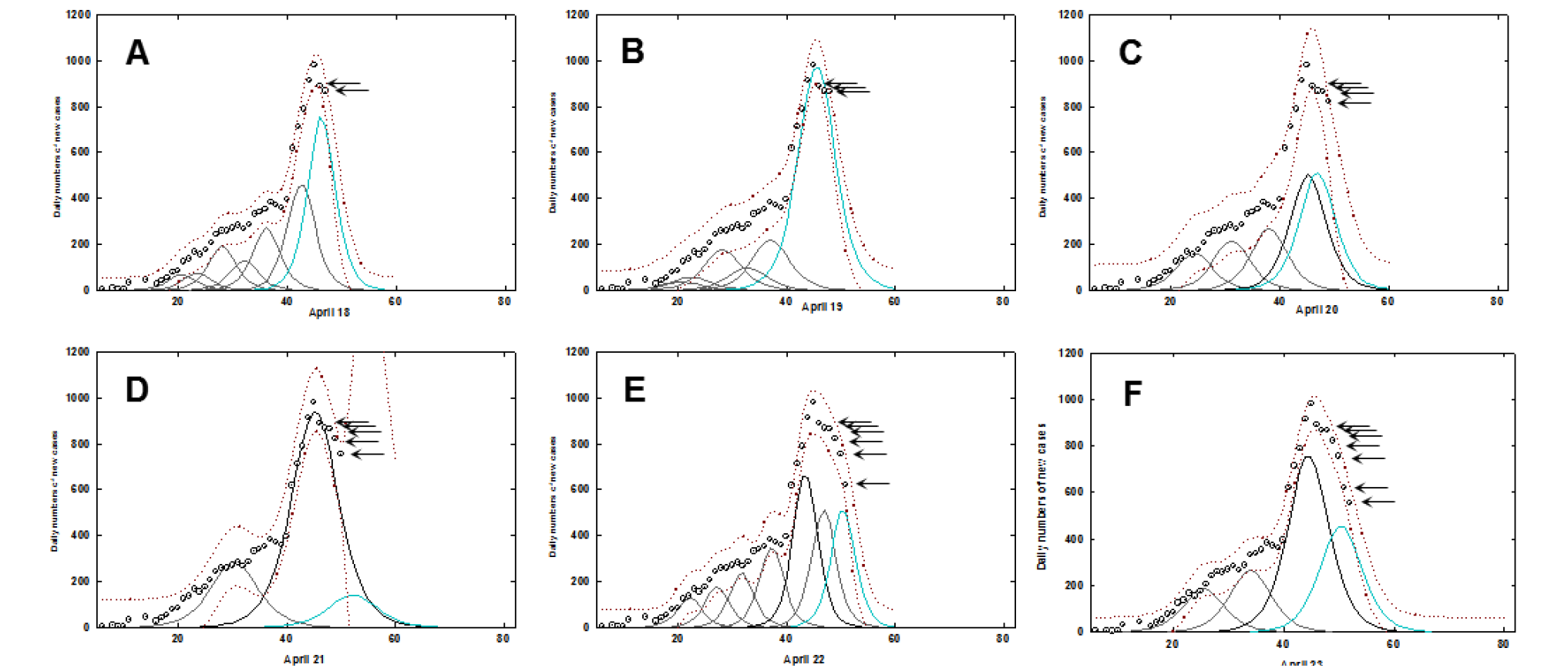
Fitted logistic curves with prediction intervals for Ireland. A-F: Individual daily time course from April 18 to April 23 for data from Ireland. Circle (5-day averaged daily confirmed infections), prediction intervals (brown dotted trace), fitted curves (black), most time recent fitted curve (cyan). Arrows indicate daily new data points. See SI Movie 1 for complete dataset.

The same analysis was applied to the datasets from the four other countries, with South Korea being the country that most closely followed the logistic progression downwards from the highest amplitude point, again illustrating an effective containment of the disease (SI Movie 2). The epidemic time progression in Iceland showed a prompt decline post peak whereas Germany had characteristics similar to Ireland. Epidemic time sequences for Germany, UK and Iceland can be viewed as supplemental Movies 3-5 respectively.

The above analysis demonstrates that variations in national scale TTQ responses have clear implications for epidemic progression in a specific region. Based on the above analysis, Ireland was median in terms of outbreak containment and as such would be a good dataset for further analysis. Daily COVID-19 infection data for HCWs in Ireland has been collected by the national authorities from the outset of the epidemic and is published on Ireland’s COVID-19 Data Hub.[11] This valuable dataset comprises all medical staff and the associated workforce within hospital or healthcare facilities and permits comparisons between HCWs and the population as a whole. The relationship between the national population and HCWs can be viewed as a set and subset where HCWs are entirely contained within the population set i.e. HCW ⊆ national population. It could be anticipated that from the initial outset of the epidemic, infections would originate in both set and subset but would quickly accumulate more in the HCW subset as patients arrive into hospitals and other healthcare settings (Figure 4). As infection numbers grow in the community, this would be reflected in the HCW numbers too. Following the peak of the epidemic wave, as infection numbers recede within the community, likewise they should do so in HCWs.

**Figure 4.**
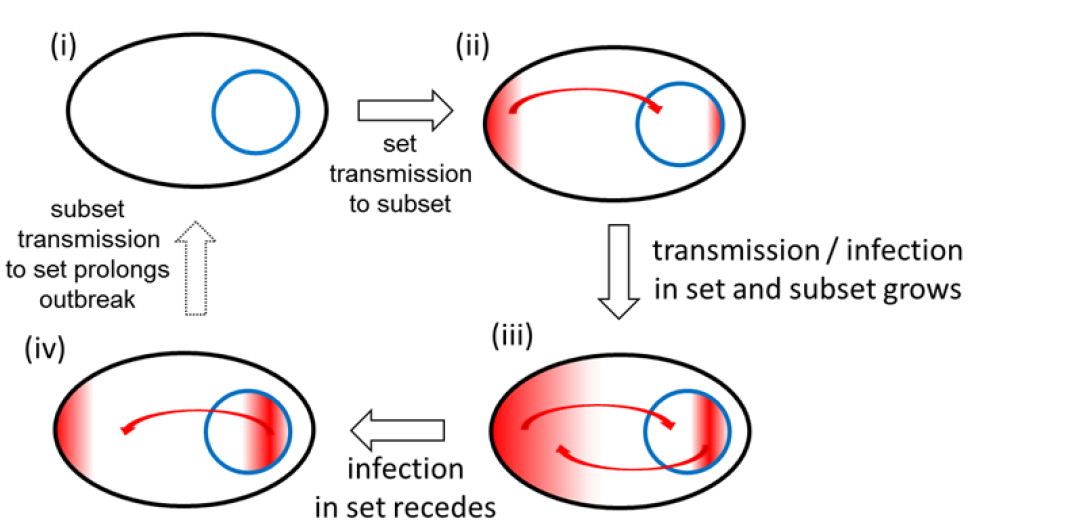
HCWs as a microcosm of society during an epidemic. Inter-relationship between set (national population) and subset (HCWs) during different pandemic phases. Infections indicated by red colour.

If the HCW population subset behaved as a relatable microcosm of the national population set during each phase of an epidemic, this raises the possibility that exhaustive testing of all HCWs, regardless of symptom status, may provide sufficient information to gauge the extent of infection in the general population. For this to be plausible, the set and subset should be consistently analogous through the growth, peak and decline phases of the epidemic. As such, a comparison of the temporal trends of HCW and entire population statistics was carried out and a correlation made as to what level HCW infection data influenced national statistics.

The number of people working in health care in Ireland has been estimated to be 119,000 with 67,000 public sector employees.[17] The collated HCW infection data was broad in scope and included all staff associated with healthcare facilities (not just medical staff) and so a benchmark number of 150,000 has been chosen to represent the HCW subset population, which is 3% of the national 4.9 M national population of Ireland. This study used 57 consecutive days of published data from March 23^rd^ to May 18^th^.[11] In absolute numbers for this time-period 7,615 of 24,036 COVID-19 positive infections or 31.6% were associated with HCWs. Figure 5A shows the very significant contribution of HCW infections to the overall total, though cumulative infection data curves of this type offer insufficient detail as to how they interrelate with each other. Analysis of the five-day rolling average daily infection numbers shows that HCWs contributed from a lowest of 19.5% to a highest of 68.9% [18] of COVID infections with an overall daily average of 33.7% throughout the epidemic (Figure 5, right). It is noteworthy that the percentage contribution generally increased as the epidemic progressed, reaching its highest as the cases in the community began to subside. This shows that despite the numerically small population of the HCW subset relative to the overall population set, its contribution to the number of COVID-19 infections is significant at all stages of the epidemic.

**Figure 5.**
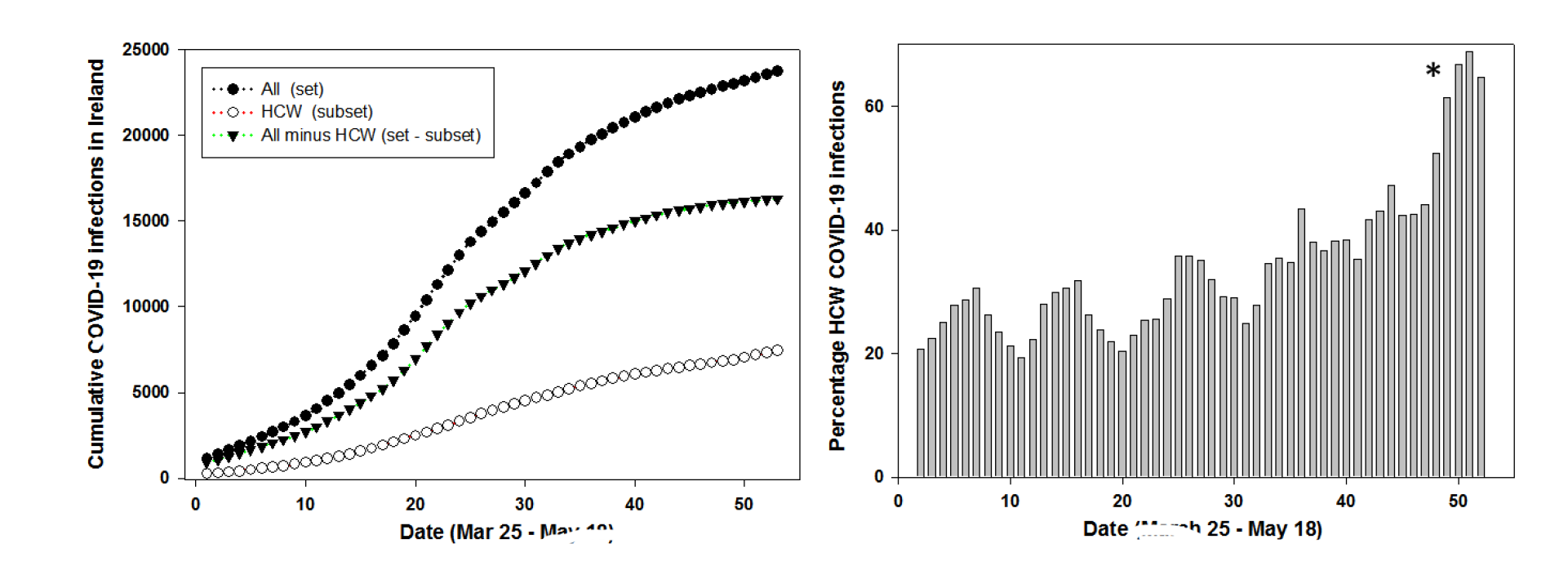
Left: Cumulative daily COVID-19 infections for Ireland (set, black circles), HCWs (subset, open circles), all infections minus HCW infections (set – subset, triangles) between Mar 25 - May 18 in Ireland. Right: Daily percentage contribution of the HCW subset to the set of all infections between Mar 25 - May 18 in Ireland. * Reference 18.

Examination of the changes in COVID-19 cases over the 57-day epidemic timespan was also revealing. Comparing the rolling averaged daily infection numbers for the entire population set with HCWs shows that they move in sequence together, mirroring each other through each of the growth, peak and decline phases (Figure 6, black circles and open circle plots).

**Figure 6.**
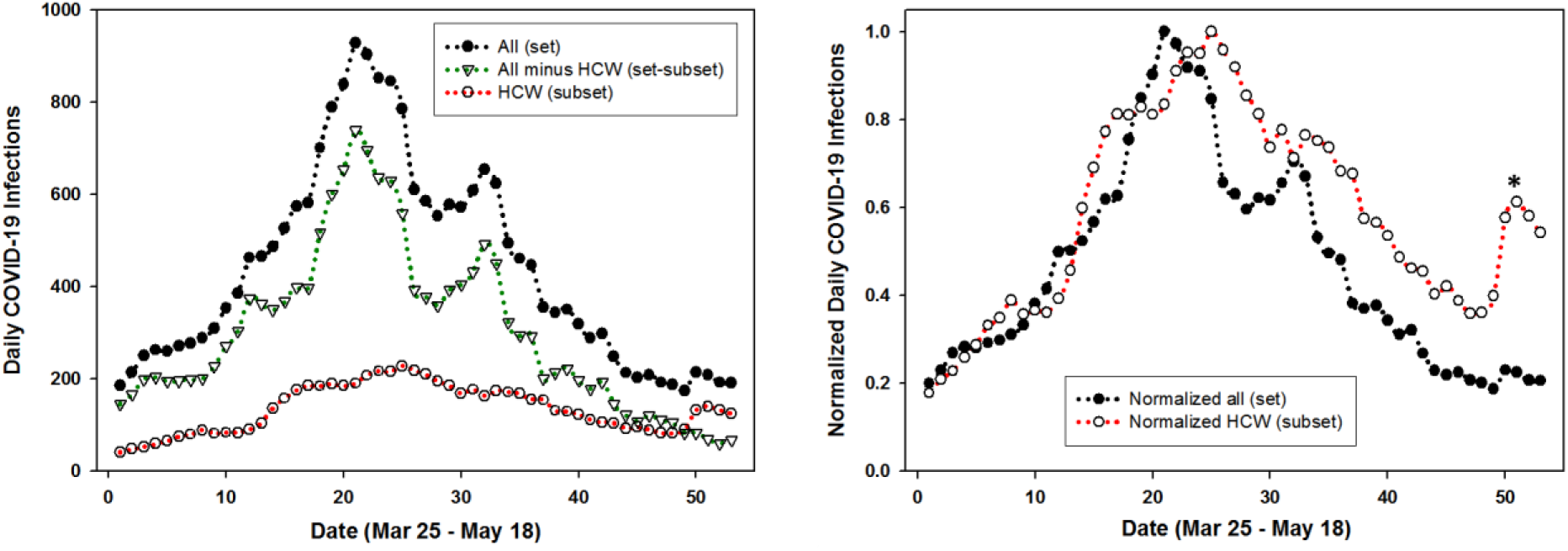
Left: Plot of five-day averaged daily COVID-19 infections for entire population (set, solid circles); HCWs (subset open circles); and entire population minus HCWs (set – subset, triangles). Right: Plot of normalized five-day averaged daily COVID-19 infections for entire population (set, solid circles) and HCWs (subset open circles) between Mar 25 - May 18 in Ireland. * Reference 18.

This is most apparent in Figure 6 (right) which shows the plot of normalized data for the entire population and HCWs indicating the potential for correlation between the set and subset. For completion, graphed data for the set minus subset is also shown (Figure 6 left, triangle labelled plot) demonstrating that excluding HCW data from the national data does not influence the trends over time.

Changes of infection growth rates over time for different population sectors is a useful comparator to determine if they have similar or interrelated patterns specifically at inflection points such as when the growth rate transitions below/above 0%. This point is an important benchmark as if a growth rate below 0% is sustained, disease transmission should diminish and ultimately stop. Comparison of growth rates of daily infections for all cases in the population with that of HCWs was revealing. Evaluation of set and subset growth factors showed similar trends for the growth phase (> 0%) of the outbreak from day 1 to 20 (Figure 7). Both set and subset transitioned from above to below 0% concurrently between days 20-30 and broadly remained below 0% from that point onwards. The growth phase trends were also similar for HCW and the entire population minus HCWs, but beyond day 25 the trend for the entire population minus HCWs was more strongly negative than that for the HCWs. These trends confirm the similarity between the set and subset and that HCWs are a higher risk subset within the entire population set.

**Figure 7.**
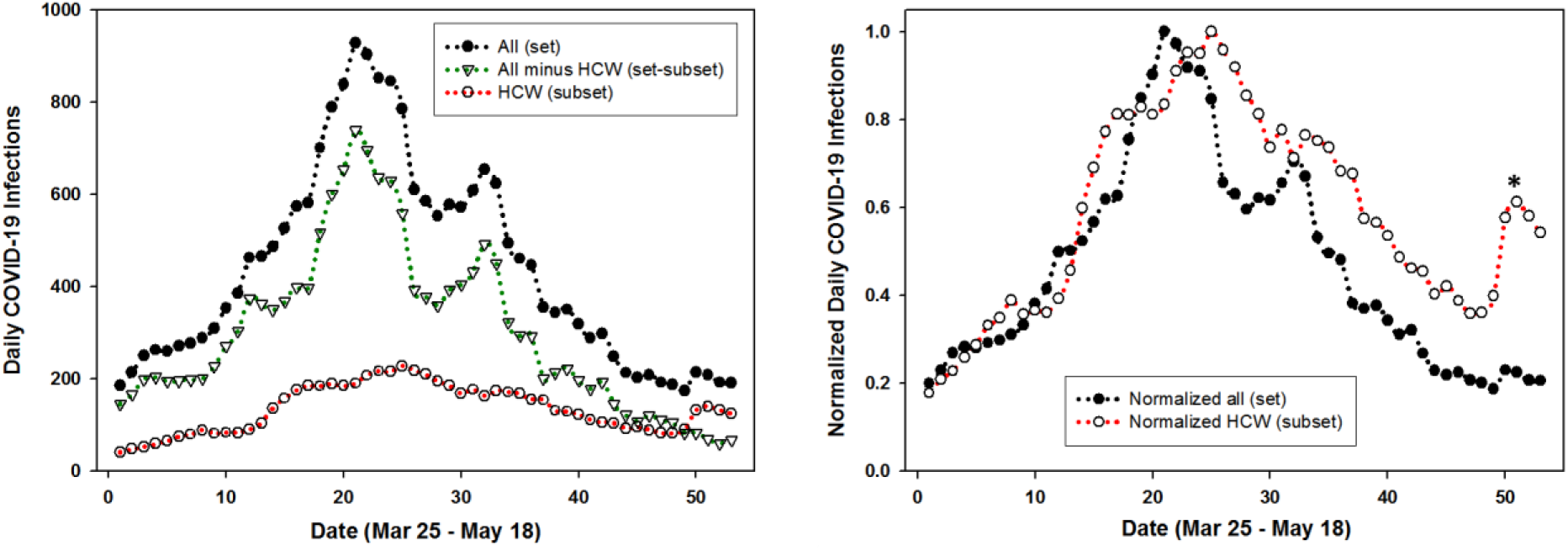
Left: Plot of the five-day moving average growth rate for all population (set, black trace) versus HCWs (subset, red trace) for Mar 25 - May 18. Right: Plot of the growth rate of all population minus HCWs (set – subset, green trace) versus HCWs (subset, red trace) for Mar 25 - May 18 in Ireland. * Reference 18.

The statistical correlation between set and subset were quantified by calculating Spearman and Pearson’s correlation coefficient rho (ρ) values. Using daily COVID-19 infection data as shown in Figure 6 coefficients for the three variants of {set / subset}, {set minus subset / subset}, and {set / set-subset} were determined (Table S1-S6). A strong correlation was obtained for the set and subset with Spearman and Pearson correlation coefficient p values of .81 and .84 respectively (Table 2). Even the more challenging correlation of set minus subset (removing HCWs from the population data) versus subset gave moderately strong correlations rho values of .71 and .74. As confirmation that these values were consistent with expectations, the values for set versus set minus subset were also determined and found to give the highest correlations. Taken together, these correlation coefficients are solid indicators that the temporal variations that occur in an entire population and its HCWs are statistically relatable.

**Table 2.**
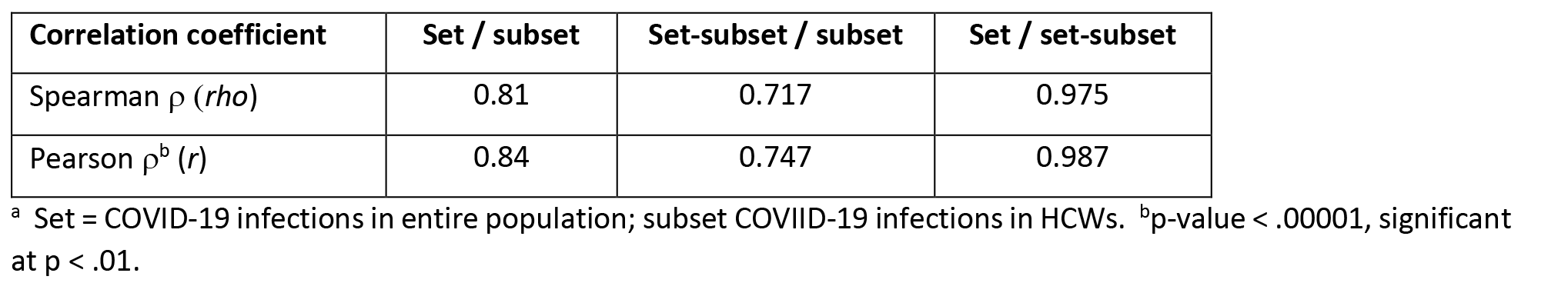
Statistical analysis of daily COVID-19 infection data for Ireland as set and subset^a^

| Correlation coefficient | Set / subset | Set-subset / subset | Set / set-subset |
| --- | --- | --- | --- |
| Spearman $\rho$ (*rho*) | 0.81 | 0.717 | 0.975 |
| Pearson $\rho^b$ (*r*) | 0.84 | 0.747 | 0.987 |

<sup>a</sup> Set = COVID-19 infections in entire population; subset COVID-19 infections in HCWs. <sup>b</sup>p-value < .00001, significant at $p < .01$ .

## Discussion

Curve segmentation of daily COVID-19 infection data has been computed and analyzed for five representative Global North countries from Europe and Asia. Operationally, smoothing algorithms were applied to averaged daily infection numbers to generate a continuous epidemic profile line for each country’s dataset (Figures 1). The area under the profile lines were fitted using sequential logistic curves which allowed cross comparison of key features of each country’s epidemic profile (Figure 2). It was observed that daily infections in countries with experienced TTQ systems efficiently declined from their maximum, thereby shortening the overall time span of their first outbreak. In contrast, countries who struggled to implement such a system experienced a more prolonged outbreak (Table 1). As this analysis can map the decline in disease transmission after the peak, it may be useful in planning and monitoring the easing of population restrictions to limit the risk of disease reemergence. To simulate its use in real-time, 95% prediction intervals were calculated following addition of each new daily data point. When deviation from the trajectory of the fitted logistic curve occurred, due to emergences of inconsistent number of infections, this was signaled by breaking of the prediction intervals. This warns that the new data is divergent from the most time forward logistic curve and that it is not accurately reflecting the epidemic at that moment in time. Consequently, an additional fitted curve is computationally added to match the emerging data as the epidemic is set to continue. Monitoring the prediction intervals for this new curve allows the process to repeat, allowing the disease progression to be tracked continually (supplementary Movies 1 and 2 for Ireland and South Korea).

To date, achieving sufficient viral testing within the general population with timely isolation of positive cases and their contacts has proved not possible for many countries. In the situation where TTQ infrastructures are insufficient to be applied to the whole population, universal testing of HCWs regardless of symptom status may act as a suitable surrogate or adjunct.[19] This hypothesis was framed mathematically using set and subset descriptors where the set is the whole population and the subset is HCWs. Using data for national and HCW COVID-19 infections in Ireland, the temporal characteristics of both were compared over 8 weeks of the epidemic. The overall statistics show that approximately 32% of all COVID-19 positive tests were for HCWs yet they are only approximately 3% of the population. Comparison of daily standard deviations and growth rates for set and subset over the course of the epidemic showed that they had similar profiles, suggesting that a comprehensive knowledge of the subset’s infection status may in fact be able to inform about the whole set (Figure 8). Spearman and Pearson correlation coefficients gave statistical measurements for a strong relationship of the relative movements of the set and subset variables (Table 2). A distinct practical advantage in testing HCWs is their relatively small population size localized within health care settings in comparison to the larger and more geographically dispersed general population.

**Figure 8.**
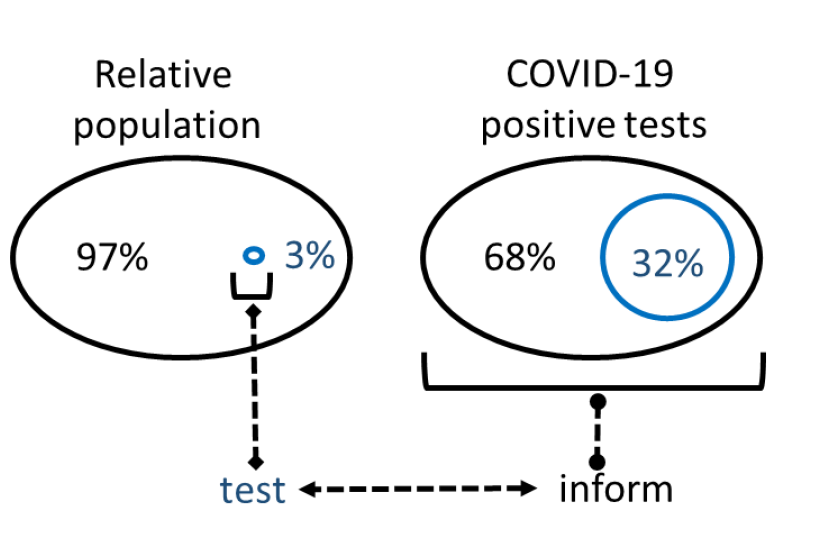
Schematic of relative populations and COVID-19 infections for the set (national population, black) and subset (HCWs, blue) in Ireland.

## Conclusion

A segmentation analysis of five country-wide epidemics has highlighted the importance of effective large scale TTQ infrastructures. Yet only a few Global North nations have been able to implement fully effective TTQ regimes, despite having the scientific and medical infrastructures to do so. Clearly, as so many Global North nations continue to fail to achieve the WHO guidance goals, nations with less medical and scientific resources will most likely be also unable to do so. An alternative that is more likely to be successfully achieved by all nations is urgently needed. A plausible approach would be to focus on a subset of the population that can inform decision making about the entire population. One such subset is HCWs, broadly defined as individuals whose employment is associated with a healthcare facility, which, due to the nature of their workplace, are known to have higher infection rates that the general community. Several practical advantages exist for continual testing of all HCWs (irrespective of symptom status) as they are more amenable to being tested, with the potential of self-testing, laboratory testing sites are often based in hospital and healthcare facilities and test sample collection could be implemented as part of the HCW’s work schedule. While not proposed as a replacement for a highly effective TTQ scheme, it may be of use if one is not available or at a minimum would augment one that is not reaching the WHO goal of test, test, test.

## Data Availability

Daily confirmed COVID-19 infection data for Germany, UK, South Korea and Iceland were taken from the open COVID-19 Data Repository provided by the Center for Systems Science and Engineering (CSSE) at Johns Hopkins University. The confirmed COVID-19 national and HCW infection data for Ireland was accessed from Ireland's COVID-19 Data Hub published by the Government of Ireland.

## Supporting Information

Movie S1 – S5 showing epidemic progression in Ireland, South Korea, Germany, UK and Iceland. Table S1-S6 Data for Spearman and Pearson coefficient determinations

## Author Contributions

### Conceptualization

Donal O’Shea

### Data curation

Pól Mac Aonghusa

### Formal analysis

Dan Wu, Pól Mac Aonghusa, Donal O’Shea

### Methodology

Dan Wu, Pól Mac Aonghusa, Donal O’Shea

### Software

Dan Wu

### Writing – original draft

Donal O’Shea

### Writing – review & editing

Dan Wu, Pól Mac Aonghusa, Donal O’Shea

